# Referral for Cardiac Amyloidosis in Patients who underwent Transcatheter Aortic Valve Replacement: Result of Quality Outcome Project

**DOI:** 10.1101/2023.06.17.23291544

**Authors:** Dae Hyun Lee, Gerry Eichelberger, Vandan Patel, Ronak Chhaya, Arjun Khadilkar, Jennifer Bishops, Hiram Bezerra, Guilherme Oliveira, Fadi Matar, Bibhu Mohanty, Joel Fernandez

**Affiliations:** Division of Cardiovascular Medicine, University of South Florida Morsani College of Medicine, Florida, USA; Department of Internal Medicine, University of South Florida Morsani College of Medicine, Florida, USA

**Keywords:** aortic stenosis, cardiac amyloidosis, quality improvement, transcutaneous aortic valve replacement, transthyretin amyloidosis

## Abstract

**Introduction:** Transthyretin cardiac amyloidosis (ATTR) is important comorbidity present in 10-15% of severe aortic stenosis (AS). The purpose of this study was to raise awareness of ATTR in patients who underwent transcatheter aortic valve replacement (TAVR) for severe AS among healthcare providers and patients.

**Methods:** We reviewed 197 consecutive TAVR cases performed from 2019-2020. Based on the presence of predefined high-risk features for ATTR based on prior literature, we subsequently contacted the patients to discuss our clinical suspicion of ATTR and offered a referral to a cardiac amyloid specialist.

**Results:** We have identified 125 patients (69.4%) who had high-risk features of ATTR. Of the 105 patients who we were able to contact, 44 patients agreed to referral, 46 patients were not able to be contacted after several attempts, and 15 patients declined referral. Of the 44 patients who agreed to referral, 20 patients completed the evaluation for cardiac amyloidosis, all of which were negative for transthyretin and light chain cardiac amyloidosis.

**Conclusion:** In conclusion, our attempt to detect ATTR in prior TAVR patients was unsuccessful after 2-3 years post-TAVR. We believe that early detection of cardiac amyloidosis close to the timing of TAVR is important and the most effective means.

## Introduction

Aortic stenosis (AS) is a common valvular disorder mostly affecting the elderly population [1]. It is caused by degenerative calcium deposition of the aortic valve. AS presents as dyspnea, chest pain, syncope, and congestive heart failure symptoms due to the pressure overload leading to left ventricle (LV) remodeling and impairment of systolic or diastolic function [1]. A relatively common disease entity that occurs in AS is cardiac amyloidosis (CA). Cardiac amyloidosis has two major subtypes: transthyretin cardiac amyloidosis (ATTR) and light-chain amyloidosis.

Multiple prospective studies have shown that CA is quite common, with about 15-20% of patients with AS having concomitant ATTR [2, 3, 4]. The pathophysiology of CA in aortic stenosis is unclear but thought to be related to high stress/ inflammation. The treatment for severe AS is surgical or transcatheter aortic valve replacement (TAVR), which improves survival [5]. Concomitant CA and AS has worse outcomes than AS without CA in meta-analysis and leading amyloidosis centers [6]{Scully, 2020 #1798}.

Both severe AS and CA can lead to heart failure. Also, severe AS and CA have some common features of echocardiogram, including cardiac hypertrophy and diastolic dysfunction. However, CA has characteristic apical sparing of strain on echocardiogram [7, 8]. Also, the discrepancy between the degree of the voltage of QRS and echocardiographic measurement of wall thickness (EKG-echocardiogram discrepancy) can be assessed through the voltage mass ratio, which is low in patients with CA [9]. The diagnosis of CA is through either an endomyocardial biopsy or a ^99m^ Technetium pyrophosphate (^99m^Tc-PYP) scan. The recent landmark ATTR-ACT trial showed the use of tafamidis in ATTR had a reduction in all-cause mortality and cardiovascular-related hospitalization/ quality of life and functional capacity [10].With the advent of novel therapeutic options with improved survival, the screening of cardiac amyloidosis became more clinically meaningful, as shown by recent studies investigating the presence of ATTR in carpal tunnel syndrome and aortic stenosis [11].

There is a gap in care and under recognition of ATTR-CA in patients who already underwent TAVR, as there was a paucity of knowledge prior to 2019 on the risk of ATTR in the setting of AS. Therefore, the aim of our study was to identify patients with high risk for the presence of CA in patients who underwent TAVR in our institution and to refer them to a heart failure cardiologist for further evaluation and screening for CA.

## Methods

This study is a single academic center quality improvement project involving patients diagnosed with severe AS who underwent TAVR. This study was reviewed by our institution’s Institutional Review Board and was approved for a quality improvement project (STUDY000938). There was no concern for ethical issues by implementing standard clinical care and practice to adhere to all federal and institutional regulations.

Based on prior literature and guideline for diagnostic testing for CA, we have selected pre-defined risk factors for the presence of CA (Table 1). We retrospectively reviewed the electronic medical records of patients who underwent TAVR at our institution from January 2019 to December 2020 (a total of 197 patients). Patients were designated high-risk group for the presence of CA if they had two or more of these risk factors. After a discussion with the structural cardiologists, a referral was made to an advanced heart failure cardiologist. Patients were contacted via phone to inform them of the quality improvement initiative and the purpose and plan for referral to a heart failure cardiologist. Heart failure cardiologists then saw patients who agreed to referral. After evaluation by the heart failure cardiologist, if a patient was deemed clinically appropriate for screening for CA, ^99m^Tc-PYP scan Anger camera was performed, along with screening for plasma cell dyscrasia through serum and urine electrophoresis, immunofluorescence and free light chain quantification.

**Table 1.** Clinical risk factors for the presence of cardiac amyloidosis in aortic stenosis

| Risk Factors | Present in High-Risk Patients (N=125) |
| --- | --- |
| Increased wall thickness on echocardiogram (PWD, IVS>12mm) | 81 (64.8%) |
| Low voltage on limb leads of 12 lead EKG | 20 (16.0%) |
| Low Sokolow-Lyon index <2.0mV | 64 (51.2%) |
| Low voltage mass ratio <1.5 | 44 (35.2%) |
| Diastolic dysfunction grade 2 or above | 64 (51.2%) |
| Low-flow low-gradient aortic stenosis | 6 (4.8%) |
| Atrial arrhythmia or Right or left bundle branch block | 75 (60.0%) |
| Systemic syndrome of amyloid | 9 (7.2%) |

The primary outcome of the quality improvement project was the diagnosis of CA in patients who underwent TAVR. We have compared the number of patients who underwent diagnostic testing for cardiac amyloidosis before our intervention (active screening for risk factors for CA) based on interventional cardiologists’ standard of care and after our intervention.

All data are presented as number (percentage %). Data were collected and managed using the REDCap electronic data management system hosted at the University of South Florida [12]. All statistical analyses were performed through R (version 4.0.4) [13].

## Results

A total of 197 patients underwent TAVR for severe AS from January 2019 until December 2020 (**Figure 1**). A total of 17 patients were deceased on the initial chart review. Of the patients who were alive on screening (N=180), there were 125 (69.4%) patients who had the high-risk criteria for the presence of ATTR and indication for referral to a heart failure cardiologist for evaluation of CA.

**Figure 1:**
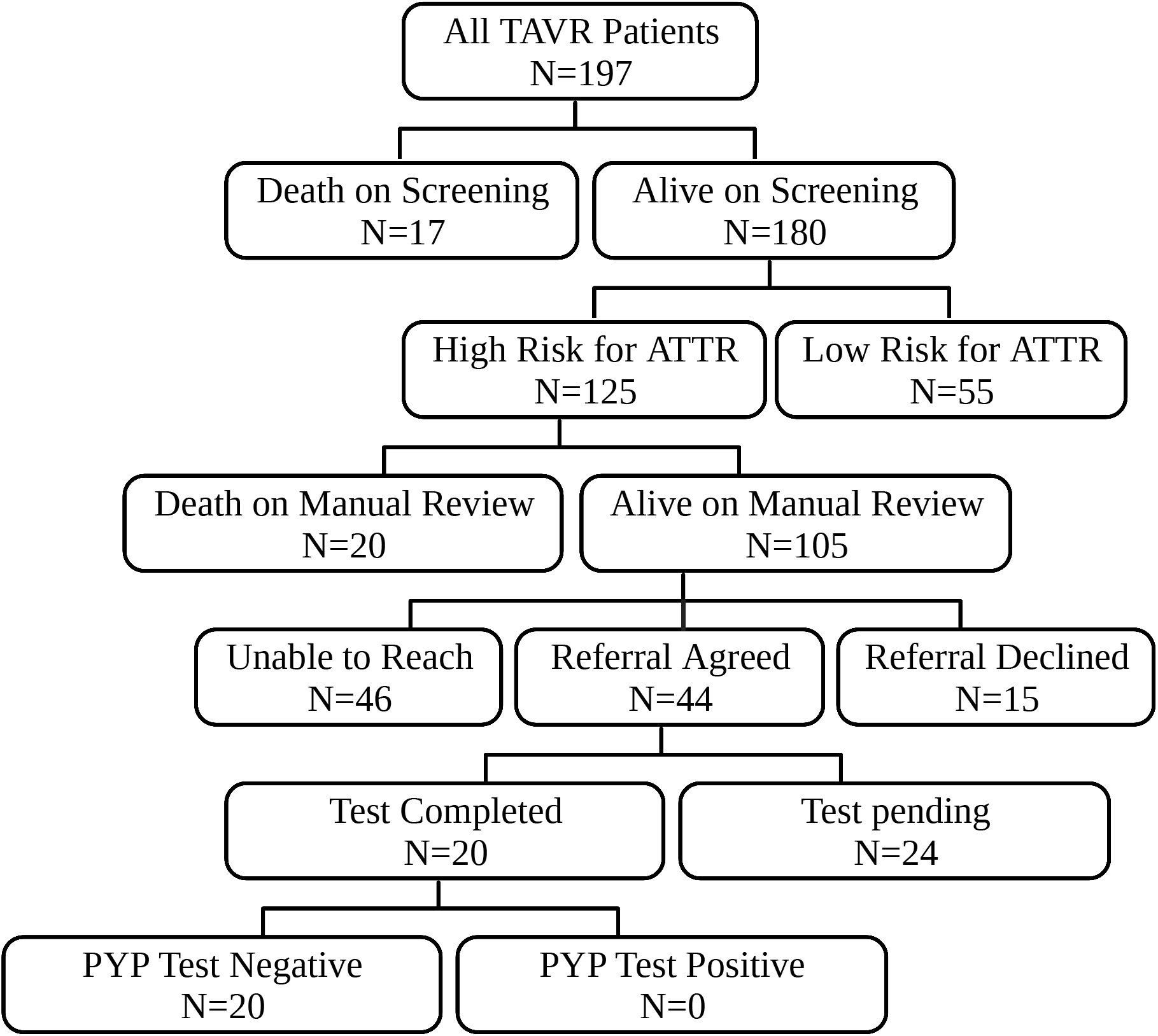
Study methodology algorithm. A total of 197 patients underwent TAVR from 2019-2020 at our institution. There were 37 patients who were deceased on chart review. There were 105 patients who had presence of high-risk features for ATTR. We attempted to contact these patients, but we were not able to contact 46 patients after multiple attempts. Of the 59 patients we were able to contact, 44 patients agreed to referral to advanced heart failure cardiologist. Twenty patients completed diagnostic tests; all were negative for ATTR or AL amyloidosis.

In patients who met the high-risk criteria for the presence of ATTR, the most common risk factors were: increased wall thickness on echocardiogram (64.8%), arrhythmias or right/ left bundle branch block (60.0%), diastolic dysfunction grade 2 or higher and low Sokolow-Lyon index (51.2%), summarized in **Table 1**.

We made attempts to contact the patients and found additional 20 patients who were deceased after the contact attempt. Of the 105 alive patients, 44 patients agreed to a referral to heart failure cardiologist, 15 patients declined the referral, and 46 patients were unable to be contacted despite three separate phone call attempts. Complete diagnostic evaluations were performed on 20 patients. However, none of the patients had a new diagnosis of CA. There are 24 patients pending complete evaluation.

## Discussion

We describe our quality improvement project on the referral of patients who underwent TAVR for evaluation of CA. Recently, multiple studies have shown the incidence of concomitant ATTR with severe AS is high at 15-20% [2, 3, 4]. The diagnosis of ATTR gained interest with the therapeutic development of tafamidis with meaningful survival benefits. After such research development, there was an increased awareness of the detection of ATTR in patients with severe aortic stenosis. However, patients who have undergone TAVR in the recent past, prior to widespread knowledge of the relation between ATTR and AS, are at risk for lack of evaluation for ATTR. Accordingly, our cohort of patients who underwent TAVR from 2019-2020 did not have a ^99m^Tc-PYP scan performed peri-procedurally.

We have utilized cardiac workups routinely performed for TAVR evaluation, including cardiac and non-cardiac history, electrocardiogram, and echocardiogram. We have utilized well-established risk factors that would now allow us to suspect for presence of cardiac amyloidosis, including low voltage on limb leads of EKG, low Sokolow-Lyon index on EKG, left ventricular hypertrophy on echocardiogram, low voltage mass index (EKG/echo discrepancy), grade 2 or higher diastolic dysfunction, presence of arrhythmia or AV/bundle branch block, low-flow low-gradient aortic stenosis, systemic syndrome of ATTR (carpal tunnel syndrome, lumbar stenosis, rotator cuff injury) [14, 15, 16]{King, 2002 #1817}. Unfortunately, at the time of our study, strain analysis was not available routinely, and we could not include apical sparing pattern as part of our criteria. A significant amount of patients (over 60%) met the high-risk criteria for the presence of cardiac amyloidosis. We initially expected that ATTR would be present in 10-15% of all patients who underwent TAVR, consistent with the literature. However, we could not identify any patients with ATTR diagnosed via PYP scan.

There are several explanations for the lack of diagnosis of cardiac amyloidosis in our cohort. First, there were a significant number of patients who we were not able to be contacted by telephone despite several attempts. To fully assess the survival of the patients we were unable to reach, we manually performed a Google search of the patient’s name and date of birth for the obituary. Second, there was a significant number of patients who were referred to heart failure clinic but not seen in the clinic/ evaluation for cardiac amyloidosis were not performed. One of the most challenging factors was the COVID-19 pandemic. This made it difficult to schedule the patients in a timely fashion. Many patients were lost to long-term follow-up since TAVR (maximum 2-3 years ago at the time of call) and the nature of tertiary referral center for TAVR. Third, the selection of high-risk features of cardiac amyloidosis may not be perfect. However, we have utilized existing literature and guidelines to decide on referral. The number of patients at high risk was much higher than the known incidence of concomitant cardiac amyloidosis and severe aortic stenosis; thus, we hypothesize that the ATTR-AS patients were all included in our high-risk patients. In that case, the ATTR-AS patients would be in the group of 1) referred but not seen, 2) deceased patients 3) unable to contact via phone. We cannot ascertain what the reason is. Lastly, a similar effort was done in patients with atrial fibrillation [17]. In the first cohort of over 800 patients with atrial fibrillation, 11.9% met the criteria for high-risk feature of ATTR, defined as age above 60 and LV wall thickness of 12mm or above. However, only 1 patient was diagnosed with ATTR. In the second cohort of patients with over 800 patients with atrial fibrillation with high-risk feature with some modification (age above 60 and LV wall thickness of 15mm or above), no patient was diagnosed with ATTR. Another limitation of our study is that we were not able to collect the cause of death in all patients, given the retrospective nature of our study. We did not collect information regarding post-mortem examination, which would have detected cardiac amyloidosis during autopsy.

Despite our effort in attempting to identify patients with concomitant cardiac amyloidosis and aortic stenosis, we were unable to identify any patients. We believe that the best screening method for cardiac amyloidosis would be at the time of evaluation for TAVR. These patients are older and may have difficulty coming to the clinic for evaluation for cardiac amyloidosis.

Therefore, evaluation for cardiac amyloidosis before TAVR during pre-TAVR evaluation or follow-up outpatient visits after TAVR may be a good time. This may be especially the case if patients have continued heart failure-like symptoms after TAVR. Further studies evaluating the screening of all or subset of patients for cardiac amyloidosis should take place, considering the cost-effectiveness and risk of extra radiation exposure from PYP scan.

In conclusion, our quality improvement project for referral of patients who underwent TAVR for evaluation of ATTR was not successful in diagnosing cardiac amyloidosis after 2-3 years after TAVR. We believe that early detection of cardiac amyloidosis close to the timing of TAVR is of importance and the most effective means.

## Data Availability

All data produced in the present study are available upon reasonable request to the authors.

## Abbreviations

AS: Aortic stenosis
ATTR: Transthyretin cardiac amyloidosis
CA: Cardiac amyloidosis
EKG: Electrocardiogram
IVS: Interventricular septum
LV: Left ventricle
PWD: Posterior wall dimension
PYP: pyrophosphate
TAVR: transcatheter aortic valve replacement
^99m^Tc-PYP: ^99m^ Technetium pyrophosphate

## Acknowledgments

The preliminary result of this project was presented at the American Heart Association Scientific Session 2021 (10.1161/circ.144.suppl_1.9413) and American College of Cardiology Scientific Session 2022 (10.1016/S0735-1097(22)01398-5). Figure 1 was created by BioRender.com

